# Smartphone App Stress Assessments: Heart Rate Variability vs Perceived Stress in a Large Group of Adults

**DOI:** 10.1101/2020.12.23.20247494

**Authors:** Konstantin Tyapochkin, Marina Kovaleva, Evgeniya Smorodnikova, Pavel Pravdin

## Abstract

**Background:** Multiple studies have shown that the state of stress has a negative impact on decision-making, the cardiovascular system, and the autonomic nervous system [1]. In light of this, we have developed a mobile application in order to assess user stress levels based on the state of their physiological systems. This assessment is based on heart rate variability [2], [3], [4], [5], which many wearable devices such as Apple Watch have learned to measure in the background. We developed a proprietary algorithm that assesses stress levels based on heart rate variability analysis, and this research paper shows that assessments positively correlate with subjective feelings of stress experienced by users.

**Objective:** The objective of this paper is to study the relationship between HRV-based physiological stress responses and Perceived Stress Questionnaire self-assessments in order to validate Welltory measurements as a tool that can be used for daily stress measurements.

**Setting:** We analyzed data from Welltory app users, which is publicly available and free of charge. The app allows users to complete the Perceived Stress Questionnaire and take heart rate variability measurements, either with Apple Watch or with their smartphone cameras.

**Subjects:** To conduct our study, we collected all questionnaire results from users between the ages of 25 and 60 who also took a heart rate variability measurement on the same day, after filling out the Questionnaire. In total, this research paper includes results from 1,471 participants (602 men and 869 women).

**Measurements:** We quantitatively measured physiological stress based on AMo, pNN50, and MedSD values, which were calculated based on sequences of RR-intervals recorded with the Welltory app. We assessed psychological stress levels based on the Perceived Stress Questionnaire (PSQ) [6], [7].

**Results:** Physiological stress reliably correlates with self-assessed psychological stress levels - low for subjects with low psychological stress levels, medium for subjects with medium psychological stress levels, and high for subjects with high psychological stress levels. On a scale of 0-100%, median physiological stress is 48.7 (95% CI of 45.2-50.7%), 56.4 (95% CI of 54.3-58.9), and 62.5 (95% CI of 59.7-66.3) for these groups, respectively.

**Conclusions:** Physiological stress response, which is calculated based on heart rate variability analysis, on average increases as psychological stress increases. Our results show that HRV measurements significantly correlate with perceived psychological stress, and can therefore be used as a stress assessment tool.

## 1. INTRODUCTION

Lazarus and Folkman define psychological stress as a type of physiological stress that arises as a result of a person’s psychological perception of life events [8]. Today, the definition “psychological stress is the process of interaction from resolution requests from the environment (known as the transactional model)” is widely accepted. [9]

Psychological stress is widely researched due to its negative effects on the human body [10]. Although moderate stress levels can be beneficial, helping individuals cope with some situations better, high psychological stress levels can have negative effects on memory, attention span, and the body’s physiological systems [11].

Psychological stress research is conducted with cognitive stress tests (Stroop Color and Word Test, math problems), in real-life scenarios (public speeches, academic exams, as well as stressful tasks such as performing surgeries), or with the help of questionnaires [6]. In this study, we use the PSQ questionnaire to assess psychological stress.

Physiological measures of stress include heart rate [2], [12], [13], heart rate variability [14], [15], [2], [16], [12], [13], [17], [18], blood pressure, electrophysiological brain responses, cortisol levels in saliva, and other measures [19], [20].

Stress impacts decision-making, and high stress has a negative impact on job performance [21]. For knowledge workers and individuals whose jobs entail decision-making, daily psychological stress assessments are important. In this study, we evaluate the accuracy of non-invasive stress assessments based on heart rate variability measurements [22] taken with the Welltory, through either the phone camera or Apple Watch.

### Objective

The objective of this paper is to study the relationship between HRV-based physiological stress responses and Perceived Stress Questionnaire stress self-assessments in order to validate Welltory measurements as a tool that can be used for daily stress measurements.

## 2. METHODS

### A. Data acquisition

RR-interval sequences from 1,471 subjects were obtained through the Welltory app, which subjects used to take heart rate variability measurements with their smartphone cameras or Apple Watch while in a resting state, after completing a stress self-assessment questionnaire (Perceived Stress Questionnaire (PSQ)) on the same day. We excluded measurements that showed possible arrhythmias in research subjects [23], as well as low-quality measurements [24], because these factors can significantly distort HRV metrics. The RR-interval sequences were used to calculate AMo, pNN50, and MedSD values.

AMo is the so-called mode amplitude presented in percent. AMo is obtained as the height of the normalised RR interval histogram (bin width 50 msec) [25], [26].

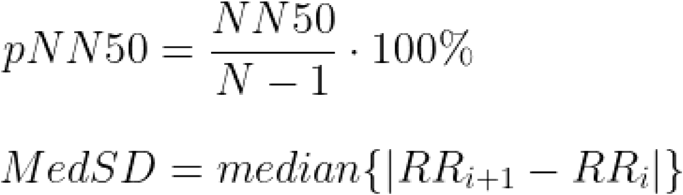

AMo and pNN50 are well-known and widely-used metrics [27], [28], [29], [30]

MedSD is similar in meaning to the widely-used metric RMSSD, but it is more robust. In order to assess psychological stress levels, we used the Perceived Stress Questionnaire (PSQ) [7], which users complete inside the Welltory app in English. This questionnaire was developed as a tool to assess stressful life events. It assesses an individual’s levels of stress across five key areas:

- measured life events
- social anxiety
- depressive symptomatology
- physical symptomatology
- perceived stress

Respondents answer questions on a scale of 1 (“almost never”) to 4 (“usually”) in order to indicate how frequently they experience specific emotions related to stress. Higher scores indicate higher stress levels.

PSQ results are presented on a scale of 30 to 120 points, and all scores are calculated in accordance with the methodology developed by the authors [6].

We used PSQ results as a benchmark for stress assessments. We split up the participants into three groups, based on their scores:

- Low stress - people with scores of 30-60 points
- Medium stress - people with scores of 61-90 points
- High stress - people with scores of over 90 points

We grouped participants based on the distribution of the total scores of PSQ (see Figure 1): the average score was 74 points with a standard deviation of 14. Thus, we defined the “medium stress” to be within the range of 60 – 90 points, that is equivalent to one standard deviation from the average. Participants with the scores below (<60) and above (>90) the cutoffs were assigned to the “low stress” and “high stress” groups, correspondingly. In general, despite some differences, our categorization principle is in line with the categorisation applied by the developers of PSQ.

**Figure 1.**
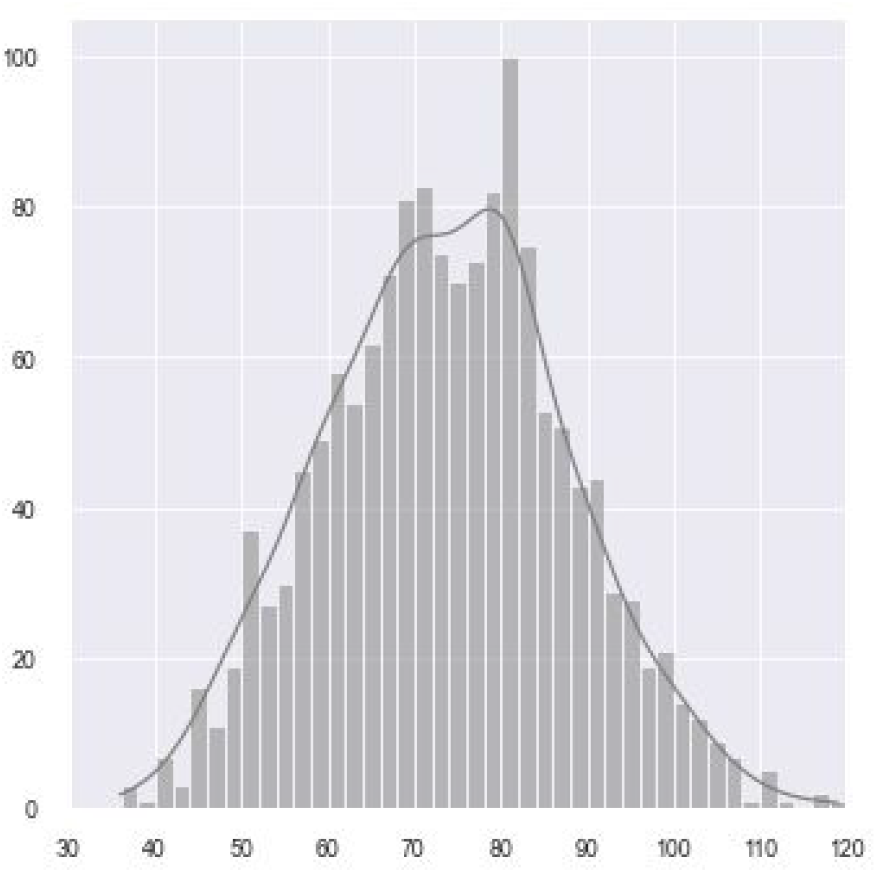
Distribution of Participant PSQ Scores

The maximum score that a participant can receive on the test, which corresponds to the highest possible stress level, is 120.

The minimum score that a participant can receive on the test, which corresponds to the lowest possible stress level, is 30.

The data was collected during the time period between 01.10.2020 and 12.12.2020 Descriptive statistics is shown in Table 1 and on Figures 2-4.

**Table 1.**
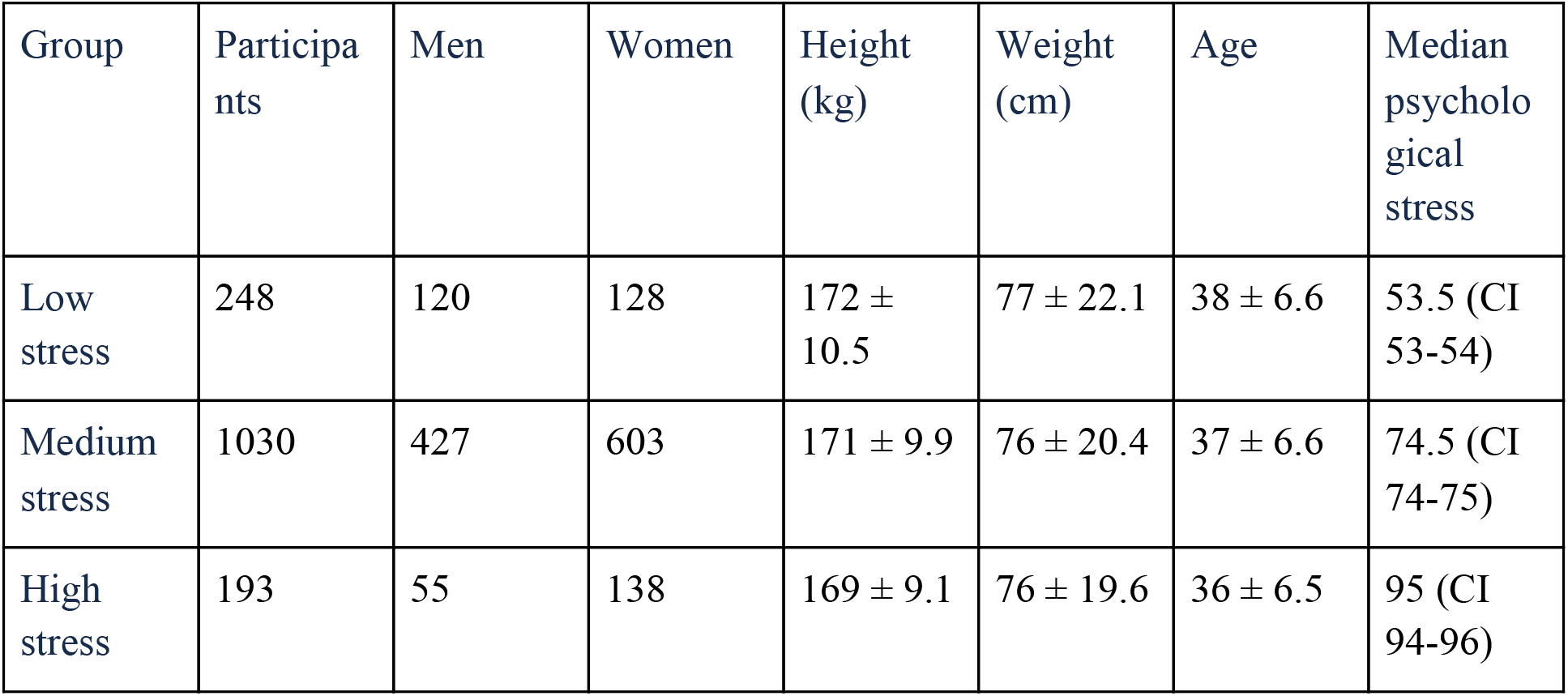
Participant Demographic Information.

| Group | Participants | Men | Women | Height (kg) | Weight (cm) | Age | Median psychological stress |
| --- | --- | --- | --- | --- | --- | --- | --- |
| Low stress | 248 | 120 | 128 | 172 ± 10.5 | 77 ± 22.1 | 38 ± 6.6 | 53.5 (CI 53-54) |
| Medium stress | 1030 | 427 | 603 | 171 ± 9.9 | 76 ± 20.4 | 37 ± 6.6 | 74.5 (CI 74-75) |
| High stress | 193 | 55 | 138 | 169 ± 9.1 | 76 ± 19.6 | 36 ± 6.5 | 95 (CI 94-96) |

**Table 2.** Median physiological stress levels for each questionnaire group.

| Group | Participants | Median stress | 95% CI |
| --- | --- | --- | --- |
| Low stress | 248 | 48.7 | 45.3 - 50.9 |
| Medium stress | 1030 | 56.4 | 54.4 - 58.9 |
| High stress | 193 | 62.5 | 59.7 - 66.3 |

**Figure 2.**
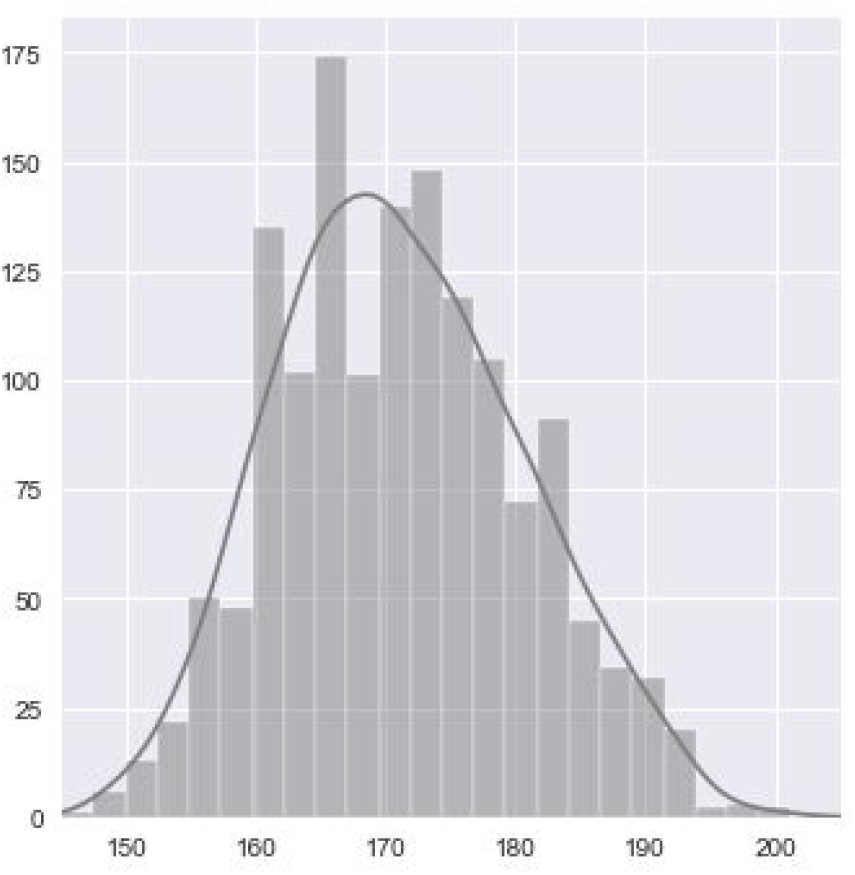
Distribution of Participant height

**Figure 3.**
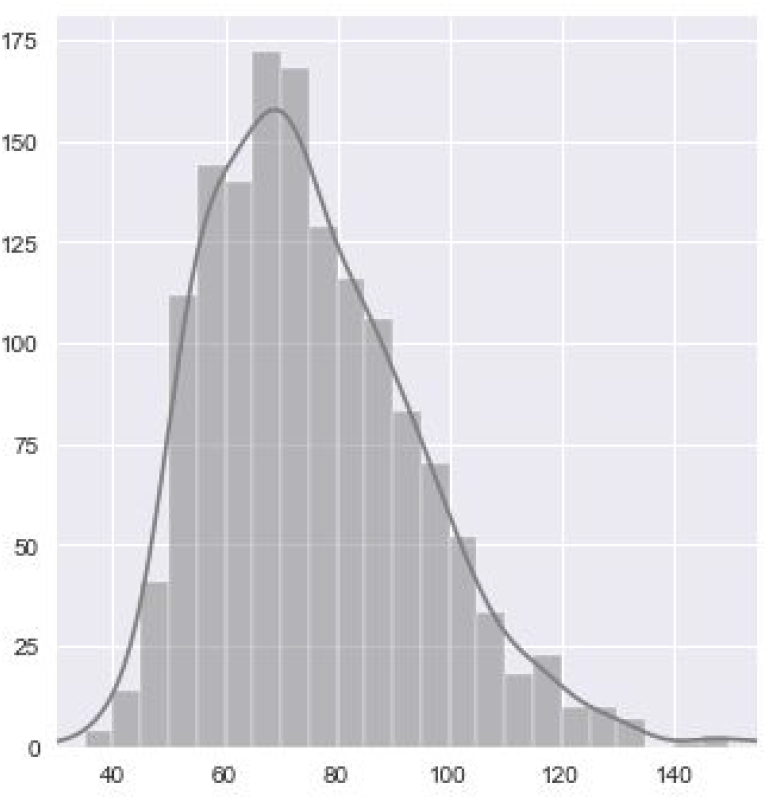
Distribution of Participant weight

**Figure 4.**
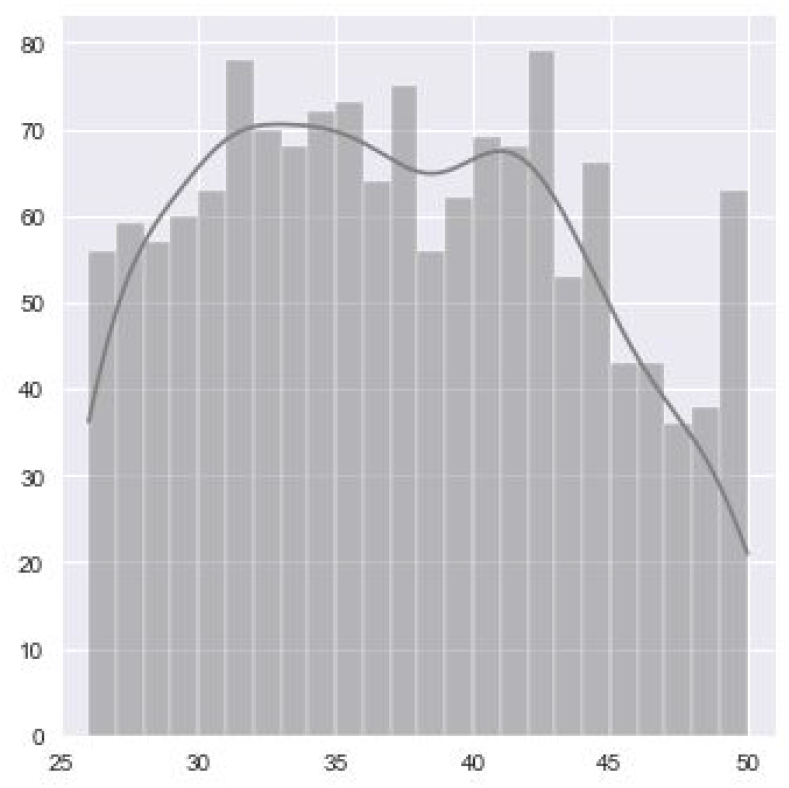
Distribution of Participant age

### B. Data Analysis

The level of physiological stress is calculated based on the deviation of AMo, pNN50, and MedSD values from the individual baselines for these values for each participant.

A percent scale was created in order to determine physiological stress levels:

- 0 - 45% - low stress. A resting state.
- 46 - 60% - optimal stress. A normal state for the individual’s regulatory systems.
- 61 - 85% - high stress. The individual’s regulatory systems are under pressure.
- 86 - 100% - very high stress. The individual’s regulatory systems are overstressed and aren’t functioning well.

The percent of physiological stress was calculated with a separate algorithm for each of the three metrics - AMo, pNN50, and MedSD. If AMo and pNN50 values were at baseline levels, this indicated optimal stress - 50%. Values that were higher than the median indicated a stress level of under 50%. The higher this deviation from the median, the lower the stress level. A drop below median pNN50 and MedSD values indicated an increase in stress. The higher this deviation from the median, the higher the stress level in %. For AMo, the reverse applied: an increase in this value above the median indicated an increase in stress above 50%, while its drop below the median indicated a decrease in stress below 50%.

After assigning a % stress value for each of these three scales, we calculated the average stress level between them. This value was then taken to indicate the level of physiological stress, in %.

### C. Study Design

This study was conducted without the active participation of the research subjects. Upon downloading the app, users provide informed consent for their anonymized data to be used by the company for internal research purposes if such research can help provide users with better services or improve the app’s functionality. This policy is described in the company’s Terms of Use, which the app’s users actively consent to.

The data is limited by the new version of the app, which was released on October 1, 2020, because the formula used to calculate stress levels based on heart rate variability was introduced in this version. Consequently, participants were only included in the study if they met the following criteria:

1. Participants filled out the Perceived Stress Questionnaire after October 1, 2020.
2. Participants took a heart rate variability measurement after completing the Perceived Stress Questionnaire.
3. Participants were between the ages of 25 and 60.
4. The quality of the heart rate variability measurements taken by participants was high enough (measurement quality is calculated based on data obtained from the measurement device [24]).

it is important to note that participants did not see a stress level based on their heart rate variability analysis prior to filling out the questionnaire. Thus, the objective physiological stress level assessment could not have had an impact on their self-assessed psychological stress level.

We only included one day of survey results in the sample, which excluded the possibility of bias due to repeated participation from the same individuals.

Thus, the data used for this study were not collected specifically for the purpose of conducting this study.

This research paper is a retrospective cross-sectional study.

The data sample includes individuals from different countries, of different ages and sexes, who took stress assessments at different times of the day.

## 3. RESULTS

In order to evaluate the results, we calculated the median physiological stress levels for each questionnaire group, along with the confidence intervals (confidence intervals were calculated with the bootstrap method [31]).

The distribution of physiological stress levels for the three groups is shown in Figure 1.

We used several approaches to compare physiological stress levels:

1. The Kruskal-Wallis H-test, or a one-way ANOVA on ranks, as a non-parametric method. H-statistic: 19.777, p-value: 0.0001
2. The Mann-Whitney U-Test for groups 1-2 and 2-3: U-statistics: 109636 and 90092, p-value: 0.0003 and 0.0194, respectively.
3. Simple Linear Regression: slope 0.191 (95% CI: 0.108 - 0.274), p-value < 0.0001

All 3 tests demonstrate that there are statistically significant differences in median values of physiological stress between these three groups, and show a positive relationship between physiological and psychological stress.

**Figure 1.**
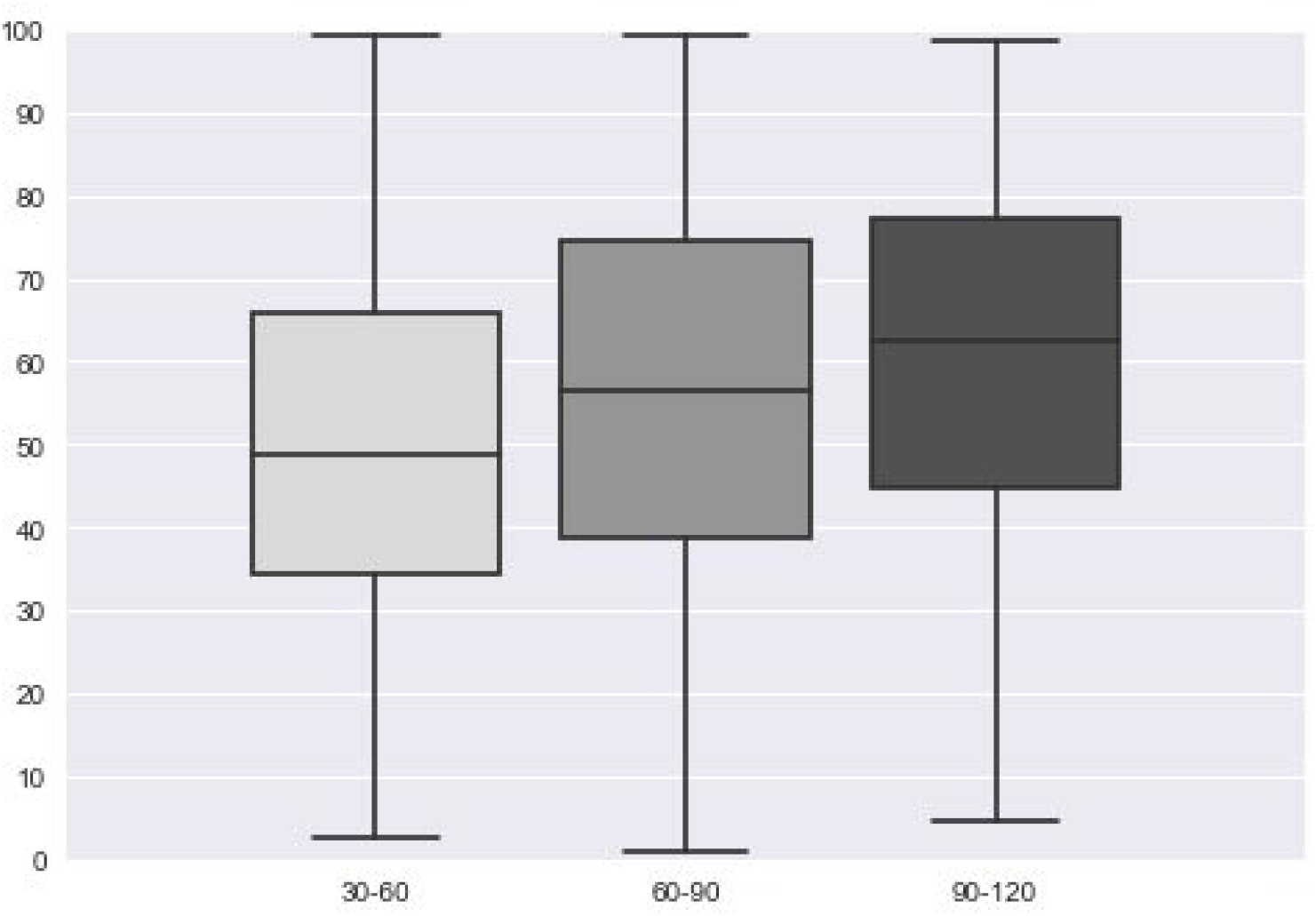
The distribution of physiological stress in low, medium, and high stress groups, as determined by the Perceived Stress Questionnaire (PSQ) results. The x-axis shows PSQ stress groups, while the y-axis shows the distribution of physiological stress levels.

**Figure 2:**
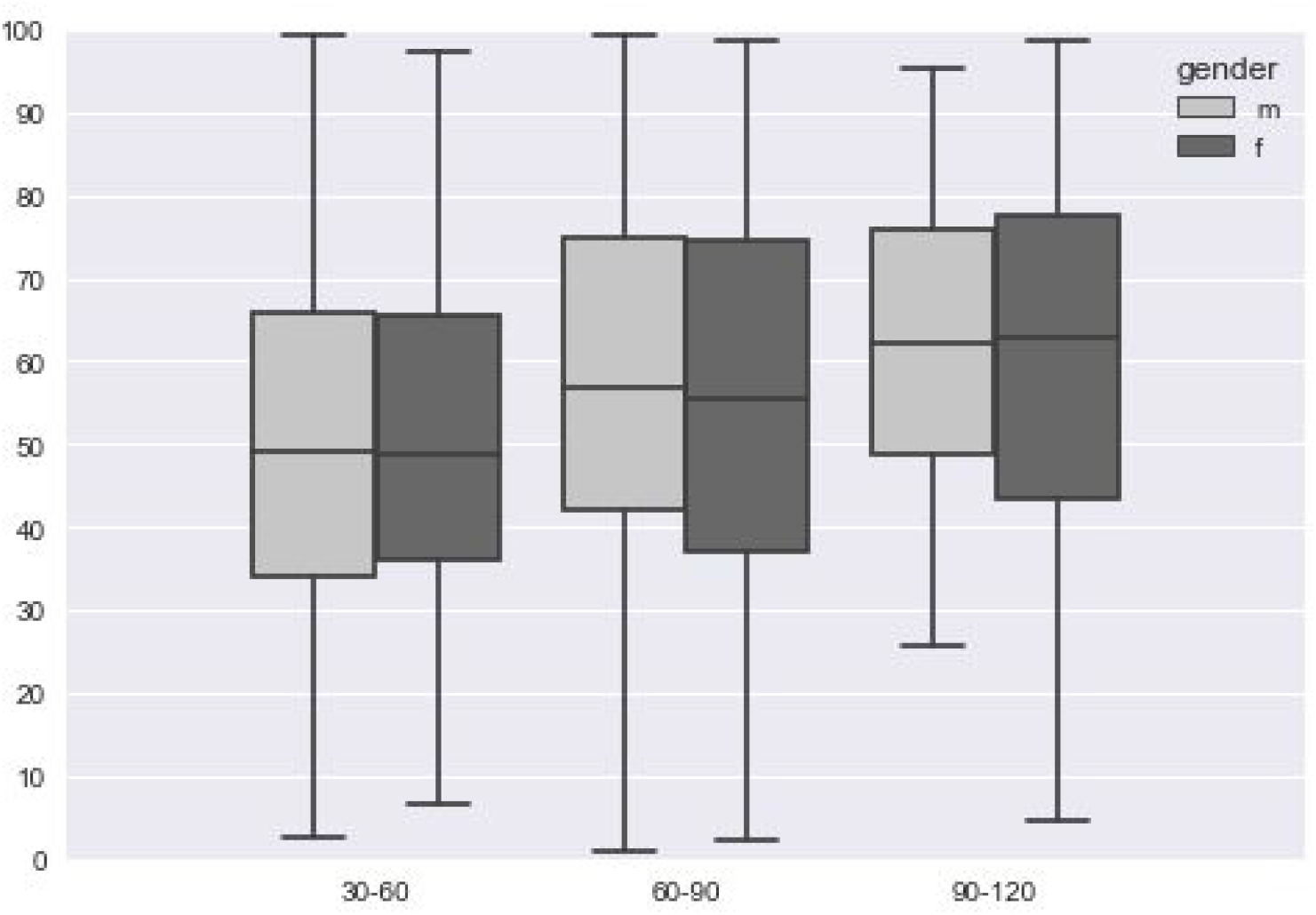
Physiological stress distribution in low, medium, and high stress groups, as determined by the Perceived Stress Questionnaire results, split by sex. The x-axis shows PSQ stress groups, while the y-axis shows the distribution of physiological stress levels.

## 4. DISCUSSION

In the beginning of the study, it turned out that there are significantly more women than men in 2 out of the 3 groups. This may have presented a potential problem in terms of verifying the results. However, we decided not to exclude the data due to the gender imbalance, but simply take it into account when analyzing the results. Figure 2 shows that there is no statistically significant difference in median stress levels between men and women (For the Mann-Whitney U-Test, the p-value > 0.05 for both men and women in each group). Thus, the imbalance in the number of men versus women did not have a significant impact on the final result.

The participants were not pre-selected in any special way: we deliberately used all data that met the criteria for the study - the study results include all individuals who downloaded the app, completed the questionnaire, and took a measurement for their own reasons. This makes the results widely applicable and increases the study’s scientific value, because stress research based on heart rate variability is typically limited by group demographics - young students [14], people in good physical shape, or people completing specific types of tasks [32], etc.

Although such studies are widely represented in scientific literature, they cannot be used to establish physiological stress norms for just any individual, because heart rate variability metrics will vary greatly depending on age, physical fitness levels, and other factors. The advantage of our approach is that it is a universally-applicable method of calculating stress levels based on comparing AMo, MedSD, and pNN50 values with baselines for different subgroups obtained from large samples. These subgroups include a diverse range of individuals, from physically fit athletes to people who lead sedentary lifestyles, as well as people of different ages.

The PSQ questionnaire we used in our study provides a subjective stress level assessment, while heart rate variability analysis measures the body’s stress response and the state of an individual’s regulatory systems. The body’s response depends on two factors: the external level of stress and stress resilience. This means that, for a resilient person in excellent health, physiological stress response may be mild even when the external stressors are significant. In such cases, Welltory will show a more objective assessment compared to self-assessment questionnaires.

However, Welltory’s stress assessment tool maybe even more valuable in the reverse scenario: when an individual’s capacity to cope with stress is low, physiological stress response may be more pronounced than perceived stress levels. This can be an important signal to reduce pressure and focus on recovery.

In this study, we established a reliable correlation between psychological and physiological stress levels. However, the variation between individual physiological stress levels was very high. This variation can be explained by the fact that, aside from psychological stress, many factors can impact the state of the body’s regulatory systems. Physical exertion and recovery, sleep, overall health, physical fitness, genetics - all of these factors impact the body. This is precisely why it was important to compare median physiological stress levels between large groups, as opposed to select individuals.

Thus, in spite of the fact that there is a positive relationship between physiological and psychological stress, physiological stress assessments are more objective - they allow individuals to accurately determine when they can continue to function under high workloads and when it is crucial to focus on recovery, regardless of what their perceived stress levels may be.

## 5. CONCLUSIONS

This study proves that physiological stress response, which is calculated based on heart rate variability analysis with a developed algorithm, on average increases as psychological stress increases. Our results show that heart rate variability measurements significantly correlate with perceived psychological stress, measured with validated PSQ inventory [6]. That’s why a developed algorithm can be used as a stress level assessment tool.

## 6. OTHER INFORMATION

The authors of this study have paid consulting agreements with Welltory Inc. The company that provided the data for this study is an interested party when it comes to the results of this research. Special official confirmation was obtained from the company, which confirms that the data provided fully matches the description of the data and were not specially selected in any way other than in accordance with the selection criteria described in this publication. The company bears no responsibility for any data modifications that may have been executed by the users but confirms that it did not prompt users to provide this data for this particular research, did not notify them that this specific data would be used for this study, did not ask for their support, and did not try to impact the received data in any other way. The company approves that necessary user consent has been obtained to conduct this research.

## Supporting information

Ethics Committee Approval Protocol

## Data Availability

Due to the nature of this research, participants of this study did not agree for their data to be shared publicly, so supporting data is not available.

## Notes

### Competing Interest Statement

The authors of this study report personal fees from consulting agreements from Welltory Inc., during the conduct of the study, as well as personal fees from Welltory Inc., outside the submitted work.

### Funding Statement

The study is supported by Welltory Inc.

### Author Declarations

The Welltory Local Ethics Committee C-003/2020 has reviewed the study named 'Smartphone App Stress Assessments: Heart Rate Variability vs Perceived Stress in a Large Group of Adults', presented by the committee secretary Yana Tatchina, on 19.12.2020 and provided their approval, signed on the Protocol #3 by December 19, 2020, attached. The study was considered the one involving no more than minimal risk to human subjects because it is based on retrospective and anonymized data. The Welltory Local Ethics Committee C-003/2020 is formed with members who do not have any affiliation with the authors, the study, or Welltory, Inc. All of the ethics committee members have declared that there is no conflict of interest. The signature pages with each approval, declaration of not having any affiliations, contacts, and research profiles of all ethics committee members are attached to this study. The list of The Welltory Local Ethics Committee C-003/2020 members who provided their approval with Protocol #3 by December 19, 2020: 1. Evgeniia Sotnikova, PhD in Mathematics, Novosibirsk State University 2. Nikolay Teslya, PhD in Software Engineering, ITMO university 3. Kseniya Solovyeva, Doctor of Philosophy, Moscow Institute of Physics and Technology 4. Tatiana Logvinenko, a Specialist degree in Clinical Psychology 5. Dmitry Repin, PhD in Cognitive Neuroscience, Boston University 6. Valery Ilinsky, a Specialist degree in biology, The European Society of Human Genetics 7. Alexei Gratchev, MD, Doctor of oncology, N.N. Blokhin Cancer Research Center 8. Andrey Perfilev, MD, an expert in personalized medicine, Atlas Biomed Group Limited

