## Supplementary material for "Smartphone App Stress Assessments: Heart Rate Variability vs Perceived Stress in a Large Group of Adults": Ethics Committee Approval Protocol

**December 19, 2020**

### Welltory Independent Research Committee Protocol #3

**Corresponding author and principal investigator:** Konstantin Tyapochkin
**Institution**: Welltory Inc., 541 Jefferson, Suite 100, Redwood City, CA 94063
**Research title**: Smartphone App Stress Assessments: Heart Rate Variability vs Perceived Stress in a Large Group of Adults, MEDRXIV/2020/247494

Dear Dr. Tyapochkin,

The Welltory Independent Research Committee has reviewed your project titled "Smartphone App Stress Assessments: Heart Rate Variability vs Perceived Stress in a Large Group of Adults," presented by the committee secretary Yana Tatchina, on 19.12.2020.

The following documents were reviewed:

a. Terms of Use & Privacy Policy for Welltory, Inc
b. Principal investigator's current CV
c. Procedures, methodology, and findings of the paper entitled "Smartphone App Stress Assessments: Heart Rate Variability vs Perceived Stress in a Large Group of Adults"

The following members of the committee were present at the meeting, held on 19.12.2020, 14:00 — 15:30 MSK.

All procedures performed in the study are in compliance with the ethical standards of the Welltory Independent Research Committee and with the 1964 Helsinki Declaration and its later amendments or comparable ethical standards. Informed consent was obtained from all individual participants whose data was used in the project. Additionally, the project meets the following criteria:

1. Is this project in conflict with the Declaration of Helsinki ethical principles? No.
2. Were participants directly involved in the research? No.
3. Can data be traced to any specific individual? No.
4. Have all participants approved the use of their data for research purposes? Yes.
5. Was this research based on retrospective and anonymized data? Yes.
6. Were the participant's data in any way provided to a third party? No.

The committee was formed with members matching the following criteria:

1. At least one expert in medicine.
2. At least one expert in biology.
3. At least one expert in mathematics.
4. At least one expert in psychology.
5. At least one expert in software engineering.
6. At least one expert in philosophy.
7. None of the experts have any affiliation with the authors or Welltory, Inc.
8. All of the experts have declared that there is no conflict of interest.

9. None of the experts were aware of the research before this review.

The committee's quorum requirements were met, and thus the members present during the meeting had the authority to make the final decision regarding the project.

An open vote was held in order to make the final decision regarding project approval. The project was approved unanimously, and the results of the vote are as follows:

| Name | Degrees | Granted approval (tap on a link to get signature page) |
|---|---|---|
| Evgeniia Sotnikova | PhD in Mathematics | Yes |
| Nikolay Teslya | PhD in Software Engineering | Yes |
| Kseniya Solovyeva | Doctor of Philosophy | Yes |
| Tatiana Logvinenko | Specialist degree in Clinical Psychology | Yes |
| Dmitry Repin | PhD in Cognitive Neuroscience | Yes |
| Valery Ilinsky | Specialist degree in biology | Yes |
| Dr. Alexei Gratchev | Doctor of oncology | Yes |
| Andrey Perfilev | MD, personalized medicine | Yes |

Summary:
Votes in favor — 8
Votes against — 0
Abstained — 0

The Welltory Independent Research Committee has reviewed your project entitled "Smartphone App Stress Assessments: Heart Rate Variability vs Perceived Stress in a Large Group of Adults," presented by the committee secretary Yana Tatchina, on 19.12.2020 and provided their approval.
The signature pages with each vote are attached to this document.

Yana Tatchina (Dec 22, 2020 13:31 GMT+3)
(signature)

Secretary, Welltory Independent Research Committee,
Yana Tatchina, PhD.

### Welltory Ethical Paper Approval Protocol #3

Final Audit Report 2020-12-22

| | |
|---|---|
| Created: | 2020-12-22 |
| By: | Evgeniya Smorodnikova |
| Status: | Signed |
| Transaction ID: | CBJCHBCAABAAAusoCk0lwxSvLUZtXbRTCseFbQ_dGq2l |

#### "Welltory Ethical Paper Approval Protocol #3" History

- Document created by Evgeniya Smorodnikova
  2020-12-22 - 10:29:30 AM GMT- IP address: 178.66.159.91

  2020-12-22 - 10:30:13 AM GMT

  2020-12-22 - 10:30:33 AM GMT- IP address: 93.158.131.181

- Document e-signed by Yana Tatchina
  Signature Date: 2020-12-22 - 10:31:31 AM GMT - Time Source: server- IP address: 5.18.242.121

- Agreement completed.
  2020-12-22 - 10:31:31 AM GMT

Adobe Sign
